# Toilets dominate environmental detection of SARS-CoV-2 virus in a hospital

**DOI:** 10.1101/2020.04.03.20052175

**Authors:** Zhen Ding, Hua Qian, Bin Xu, Ying Huang, Te Miao, Hui-Ling Yen, Shenglan Xiao, Lunbiao Cui, Xiaosong Wu, Wei Shao, Yan Song, Li Sha, Lian Zhou, Yan Xu, Baoli Zhu, Yuguo Li

## Abstract

**Background:** Respiratory and faecal aerosols play a suspected role in transmitting the SARS-CoV-2 virus. We performed extensive environmental sampling in a dedicated hospital building for Covid-19 patients in both toilet and non-toilet environments, and analysed the associated environmental factors.

**Methods:** We collected data of the Covid-19 patients. 107 surface samples, 46 air samples, two exhaled condensate samples, and two expired air samples were collected were collected within and beyond the four three-bed isolation rooms. We reviewed the environmental design of the building and the cleaning routines. We conducted field measurement of airflow and CO_2_ concentrations.

**Findings:** The 107 surface samples comprised 37 from toilets, 34 from other surfaces in isolation rooms (ventilated at 30-60 L/s), and 36 from other surfaces outside isolation rooms in the hospital. Four of these samples were positive, namely two ward door-handles, one bathroom toilet-seat cover and one bathroom door-handle; and three were weakly positive, namely one bathroom toilet seat, one bathroom washbasin tap lever and one bathroom ceiling-exhaust louvre. One of the 46 air samples was weakly positive, and this was a corridor air sample. The two exhaled condensate samples and the two expired air samples were negative.

**Interpretation:** The faecal-derived aerosols in patients’ toilets contained most of the detected SARS-CoV-2 virus in the hospital, highlighting the importance of surface and hand hygiene for intervention.

**Funding:** The work were partially supported by the National Natural Science Foundation of China (no 41977370), the Research Grants Council of Hong Kong’s (no 17202719) (no C7025-16G), and Scientific Research Fund of Jiangsu Provincial Department of Health (no S21017002).

## Introduction

The coronavirus disease 2019 (Covid-19), which is caused by the SARS-CoV-2 virus, has rapidly spread around the globe, with nearly one million confirmed cases and nearly 50,000 deaths recorded by 2 April 2020.^1^ The epidemic characteristics suggest that droplets during close contact and fomites may mediate transmission of the SARS-CoV-2 virus.^2^ It has also been ‘envisaged’ that there may be airborne spread due to certain aerosol-generating procedures in healthcare facilities.^2^ The role of the faecal-oral route remains to be determined, following the detection of the virus in stools.^3-5^ Crucially, however, the relative importance of these routes remains unknown. Significant infection has also occurred in hospitals. According to China CDC Weekly 2020,^6^ 1,716 of the 44,672 Covid-19 cases confirmed in China by 11 February 2020 were in healthcare workers. Thus, understanding the infection risk in a hospital environment is essential to protect healthcare workers.

We performed environmental sampling in four occupied isolation rooms housing 10 Covid-19 patients in The Second Hospital of Nanjing, China and analysed the association between the sampling results and the environment, as well as the transmission risk of the SARS-CoV-2 virus. The studied infectious disease hospital was built in 2015, and is now a designated hospital for receiving Covid-19 patients during the epidemic.

## Methods

### Patient data

We collected basic data of the Covid-19 patients in the sampled isolation rooms, including their date of onset of symptoms, throat-sample PCR results, CT results, symptoms, and mask-wearing behaviour. The data for these patients are summarised in Table S1. On each date, patients and their rooms, were randomly chosen for sampling. This study was part of the Jiangsu CDC’s epidemiological studies for the Covid-19 outbreak; ethical approval was waived for the study.

### Environmental sampling

Our environmental sampling was conducted in four isolation rooms, a nursing station, a corridor, an air-conditioning system, and other spaces in the airborne infectious-disease zone on the fifth floor of the hospital (Figure 1a). The hospital is a six-storey building with a courtyard. The sampling was conducted on 8, 20, and 22 February 2020. At the time of our sampling, only some of the 19 isolation rooms in the studied zone were occupied, by 34 patients on 8 February, 21 patients on 20 February, and 34 patients on 22 February. Each isolation room has three beds and measures 7·9 m × 3·9 m × 8·2 m, but it is not necessarily fully occupied.

**Figure 1.**
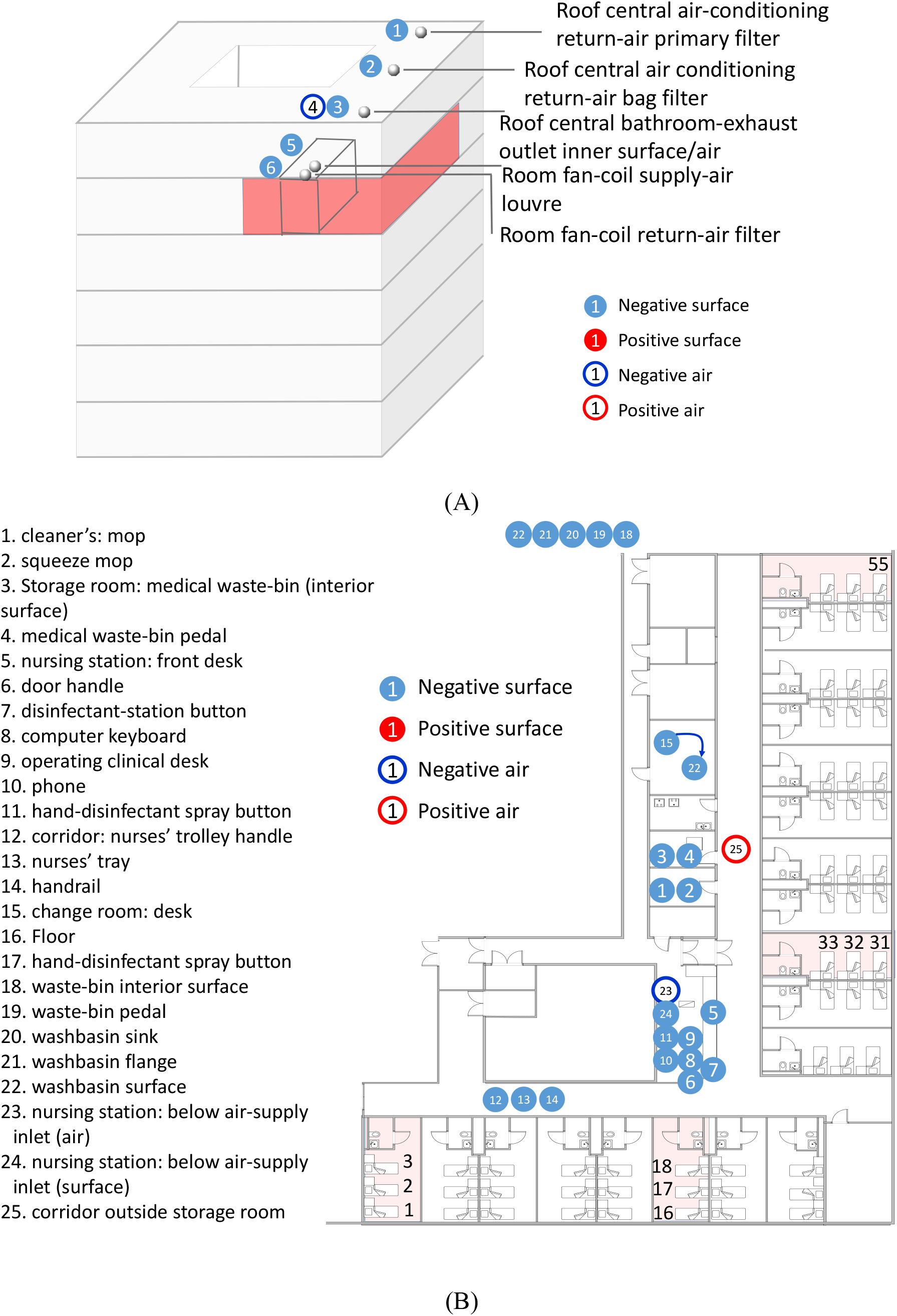
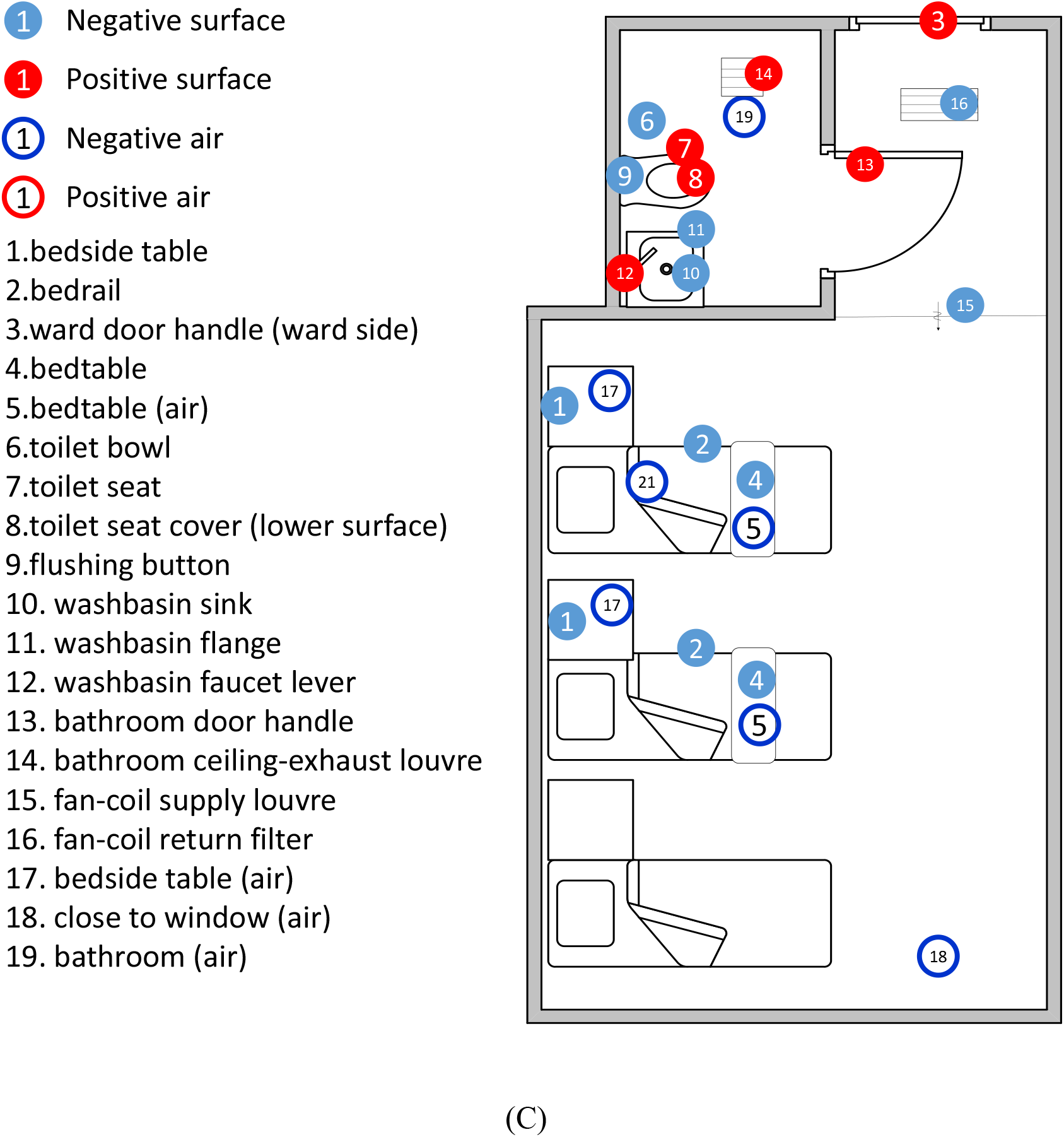
Summary of the sites where the air and surface samples were taken in the hospital. (A) In the air-conditioning systems of the isolation rooms and on the building roof. The highlighted area in red is the airborne infectious disease zone on the 5^th^ floor; (B) at the nursing station, storage/cleaner’s rooms, healthcare workers’ PPE changing room, and corridor; and (C) in the isolation rooms, but note that in some of the rooms, fewer surface and air samples were taken than are shown here. The positive samples are shown by either empty circle in red or filled circle in red. The negative samples are shown by either empty circle in blue or filled circle in blue. In Figure 1B, the four sampled rooms are highlighted in light red, i.e. isolation rooms containing beds 2 and 3, beds 16-18, beds 31 and 32, and bed 55.

**Figure 2.**
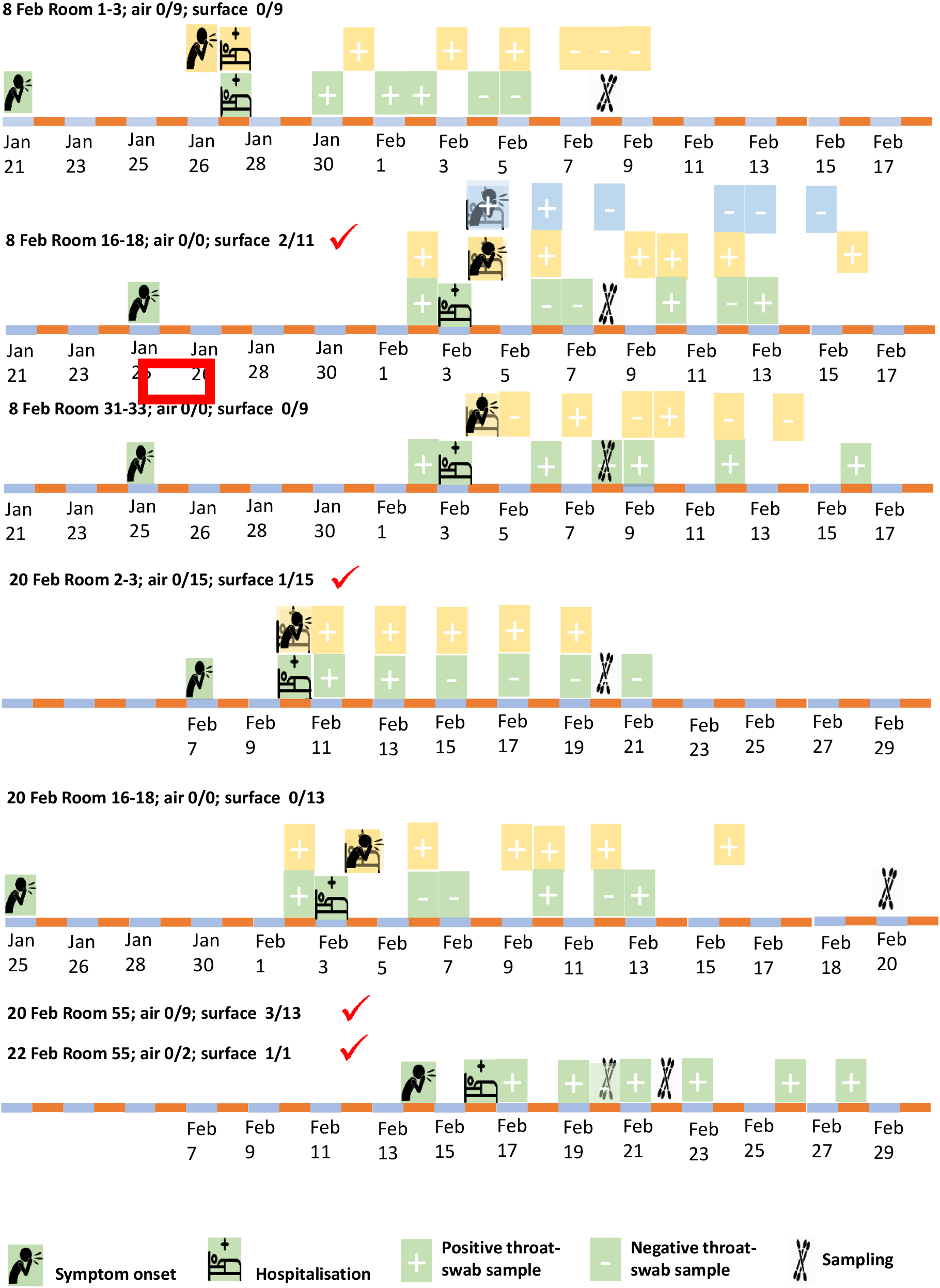
Summary of the isolation rooms containing beds 2 and 3, beds 16-18, beds 31 and 32, and bed 55, and their 10 patients (onset and hospitalisation dates), and the sampling dates. The sampling dates on which positive samples were detected are also shown by a red tick. In each room, a patient and his/her bed are shown in the same colour. When events occurred on the same day, such as symptom onset and hospitalisation, the symbols overlap and are shown in transparent style.

The sites for the air and surface samples are shown in Figure 1. All samples were collected by a trained CDC officer who had a medical background, assisted by two trained nurses from the hospital. The CDC officer wore full personal protective equipment (PPE), namely an N95 respirator, goggles, face shield, gloves, shoe covers, cap, and gown. He was quarantined for 12 days after the collection on 8 February, disinfected, and resumed sampling on 20 and 22 February. Thereafter he was quarantined again for another 14 days, and again disinfected.

For airborne-aerosol sampling, four bioaerosol samplers were used, i.e., an Andersen one-stage viable impactor (QuickTake-30, SKC, USA) (sampled at 10 L/min for 30 min, Gibco cell-culture medium, 10 mL) (only used on 8 February, as its flow rate was found to be too low), an AirPort MD8 (Sartorius, Germany) (50 L/min for 20 minutes, water-soluble gel film), an ASE-100 (Langsi Medical Technology, Shenzhen, China) on 8 February (500 L/min for 2 min, biological aerosol special-collection liquid) and on 20 and 22 February (500 L/min for 20 min, biological aerosol special-collection liquid), and a WA-15 (Dinglan Technology, Beijing, China) on 20 February (sampled at 14 L/min for 30 min, Youkang virus-sampling kit, Youkang Hengye Biotechnology, Beijing, China). For collecting exhaled breath condensate, an AT-150 (Dingblue Technology, Beijing, China) was used to obtain samples from respiratory fluids, which aggregate on the hydrophobic film’s surfaces after freezing. For surface-wiped samples, we used a Youkang virus-sampling kit (Youkang Hengye Biotechnology, Beijing, China). A cotton swab was moistened with the collection liquid, and then used to wipe the surface of the object once.

We extended our sampling sites to other areas in the hospital and its roof air-exhausts on 20 and 22 February. We also extended the sampling period of the ASE-100 from 2 min to 20 min. A detailed list of air and surface samples and sampling sites are given in Table S2 and Figure 1. Our sampling was conducted from 9:00 am to 12:30 pm, and the morning surface-cleaning in the hospital was arranged to be suspended. The usual surface-cleaning routine instead took place in the afternoon, after we had completed our sampling.

In total (Table S1), we collected 107 surface samples, 46 air samples, two exhaled condensate samples, and two exhaled air samples. On 8 February, we collected 60 samples, comprising nine air samples (with the QuickTake30, MD-8, and ASE-100) and 51 surface samples. On 20 February, we collected 29 air samples (with the MD-8, ASE-100, and WA-15), two exhaled condensate samples, two exhaled air samples, and 50 surface samples. On 22 February, we collected eight air samples (using the MD-8 and ASE-100) and six surface samples.

Environmental sample detection included nucleic acid extraction (NP968, Tianlong Science & Technology, Xi’an, China), and amplification by real-time quantitative polymerase chain reaction (RT-PCR; Applied Biosystems QuantStudio Dx (Thermo Fisher Scientific, Waltham, USA). Detection of the SARS-CoV-2 viral RNA was done using real-time RT-PCR kit from Shanghai Chromysky Medical Research Co. Ltd. (Shanghai, China). According to the instructions provided by the company, each sample was run in duplicate by RT-PCR. The patient sample was analysed by The Second Hospital of Nanjing using Applied Biosystems 7500 Real-Time PCR Systems (Thermo Fisher Scientific, Waltham, USA). The nucleic acid detection reagents were from Huada Biotechnology (Wuhan) Co., Ltd. A sample is positive when CT ≤ 38, and weak positive when 37 ≤CT ≤ 38.

We also conducted chi-squared test to determine whether there is a statistically significant difference in percentages of positive samples among different groups of surfaces. Specifically, we calculated the chi-square test statistic, X^2^, and compared the value of this statistic to a chi-squared distribution to acquire the P-value. In this study, a P-value less than 0.05 was considered to be statistically significant.

### Building and its ventilation data

We reviewed the design drawings of the building and its air-conditioning system, floor plans, and air-cleaning routes in the airborne infectious-disease zone. We conducted field measurements on 5 March in one of the identical wards on the fourth floor. We monitored the airflow direction through doorways and the bathroom door using smoke visualisation. In the area comprising the supply outlet, return inlet, and exhaust inlet in the bathroom, we measured the air speed using an anemometer (425, Testo, Germany), and recorded measurements as average values. We measured CO_2_ concentrations in the rooms, with four people present, using an indoor air-quality meter (IAQ-Calc 7515, TSI, USA) for 24 min. We investigated the airflow pattern in the room using incense smoke, with no patients present. Finally, we estimated the ventilation rates from the measured CO_2_ concentrations and air speeds.

### Role of the funding source

The funding bodies had no such involvement in study design; in the collection, analysis, and interpretation of data; in the writing of the report; and in the decision to submit the paper for publication.

The corresponding authors also confirms that they had full access to all the data in the study and had final responsibility for the decision to submit for publication.

## Results

The studied hospital is one of the dedicated Covid-19 patient hospitals in Nanjing. The hospital is a six-storey building, and our sampled area, patient zone no. 5, occupies part of the fifth floor (Figure 1a). Four patient rooms were monitored, and sampling was also conducted in the corridor and elsewhere on the floor and on the hospital roof. The sixth-floor wards have an anteroom, but fifth-floor wards, where we performed environmental sampling, do not.

The 5^th^ floor has a fan coil air-conditioning system with an outdoor-air supply. Two fans deliver 1000 m^3^/h and 8500 m^3^/h (2300 L/s) outdoor air to the non-patient and patient rooms, respectively. No ventilation is provided to the corridor, and a positive pressure is maintained at the nursing station, with no exhaust. According to the original design, each of the 19 isolation rooms is provided with an average of 47 L/s of outdoor air (2.0 ACH). The exhaust-air fan in the bathroom of each isolation room operates continuously, creating a negative pressure in the ward. Based on the CO_2_ data, the outdoor air-supply rate was calculated to be 64 L/s (2.5 ACH). The airflow pattern was found to lead from the corridor to each isolation room, and to subsequently exhaust via the bathroom.

Cleaning and disinfection in these rooms was conducted twice daily. The frequently touched surfaces were cleaned using sodium dichloroisocyanurate solution containing 500 mg/L chlorine (disinfectant tablets, Lvshaxin Aiershi, Shanghai). During the morning round, cleaners changed clothes to PPE and entered at 8:30, emptied the waste bins at 8:30-9:00, cleaned environment surfaces from 9:00 to 10:30, and exited at 11:00. During the afternoon round, cleaners entered at 2:00, and cleaned the same surfaces at 2:30-4:00.

Of the 46 air samples, 45 were negative. The only weakly positive air sample was obtained from the corridor close to the patients’ isolation rooms, and was collected on 20 February using the ASE-100 with a total air-volume of 10 m^3^. Of the two exhaled condensate samples, both were negative. Of the 107 surface samples, seven were positive. These samples were from the inside door-handle of the isolation room containing beds 16, 17, and 18 (CT = 36.8, 407 RNA copies, 8 Feb), the toilet seat in the same isolation room (CT = 38.0, 8 Feb), the inside door-handle of the isolation room containing beds 2 and 3 (CT =36.2, 666 RNA copies, 20 Feb), the toilet-seat cover (lower surface) in the isolation room containing bed 55 (CT = 36.1, 723 RNA copies or 29 copies/cm^2^, 20 Feb), the bathroom tap-lever of the same room (CT = 37.7, 20 Feb), the bathroom door-handle of the same room (CT = 36.8, 407 RNA copies, 20 Feb), and the exhaust air-grille surface in the bathroom of the same room (CT = 37.9, 22 Feb). Note that we did not calculate the RNA copies for the three weakly positive samples.

## Discussion

Among the 107 surface samples taken throughout and beyond the infectious disease zone, four were positive and three were weakly positive. Five of the seven positive/weakly positive samples were obtained from two bathrooms used by patients, with all the throat swabs of at least one of these patients having tested positive for two days before sampling. In addition, among all other surface areas both within and beyond the isolation rooms, two surface samples were positive, namely the door handles of Rooms 16-18 and 2-3.

The results suggest that for the Covid-19 patients studied here, the toilet is the most contaminated environment, although the chi-square *P* value is only 0·064 (Table 2). Our detection of more positive surface samples in the bathroom suggests that these samples may be faecal in origin. The previous detection of the virus in stools^3-5^ provides support for this interpretation, as does the fact that stools obtained from the first Covid-19 patient in the United States also tested positive.^7^ Zhang et al.^5^ also found that there were more anal swab positives than oral swab positives in the later stage of infection, and diarrhoea and nausea prior to fever and dyspnoea also presented in approximately 10% of the patients in Wuhan.^8^ High levels of viral load have also been detected in stool in SARS-CoV patients.^9^

**Table 1.** Summary of samples collected on the three sampling dates in the four isolation rooms and other areas in the airborne infectious-disease zone. The rooms containing a patient whose throat swabs were all positive on a given day (Table S1) are shown in red. The numbers of air samples and surface samples that are positive, and the corresponding shortest symptom onsets in the sampling period, are also shown.

| Sampling date and time | Locations | Indoor air temperature and humidity | Occupancy (no. of patients) | Air samples (positive/total) | Surface samples (positive/total) | Days post-symptom onset | Main objects or surfaces |
| --- | --- | --- | --- | --- | --- | --- | --- |
| 8 Feb<br>10:00am – 1:30 pm<br>Last cleaning<br>2:30–4:00 pm 7 Feb | Room 1-3 | 19–20°C<br>28–29% | 2 | 0/9 | 0/9 | 13 |  |
|  | Room 16-18 |  | 3 | 0/0 | 2/11 | 4 | Entrance-door handle and toilet seat |
|  | Room 31-33 |  | 2 | 0/0 | 0/9 | 4 |  |
|  | Other areas (cleaner's storage, nursing station, and PPE exchange) |  | NA | 0/0 | 0/22 | NA |  |
| 20 Feb<br>1:30pm – 5:30 pm<br>Last cleaning<br>2:30–4:00 pm 19 Feb | Room 1-3 | 21–23°C<br>27–29% | 2 | 0/15 | 1/15 | 10 | Entrance-door handle |
|  | Room 16-18 |  | 2 | 0/0 | 0/13 | 16 |  |
|  | Room 55 |  | 1 | 0/9 | 3/13 | 6 | Entrance-door handle, toilet bowl, washbasin spout |
|  | Roof exhaust/return | NA | NA | 0/2 | 0/5 | NA |  |
|  | Other areas |  | NA | 1/3 | 0/4 | NA | Corridor air |
|  | Exhaled condensate |  | NA | 0/2 | NA | NA |  |
|  | Exhaled air |  | NA | 0/2 | NA | NA |  |
| 22 Feb<br>11:00am – 4:30 pm<br>Last cleaning<br>2:30–4:00 pm 21 Feb | Room 55 | 22–23°C<br>33–34% | 1 | 0/2 | 1/1 | 8 | Toilet-exhaust louvre |
|  | Other areas | NA | NA | 0/6 | 0/5 | NA |  |

**Table 2.** Statistical significance of the positive results for toilet-related surfaces or surfaces in isolation rooms containing at least one patient who had all-positive throat swabs before and/or after the sampling.

| Surface groups | Number of samples | Number of positive and weakly positive samples (percentage) | *P* value (chi-square) |
| --- | --- | --- | --- |
| Toilet-related surfaces | 37 | 5 (13.5%) | 0.064 |
| Other surfaces within the isolation rooms | 34 | 2 (5.9%) |  |
| Other surfaces beyond | 36 | 0 (0%) |  |

| the isolation rooms |  |  |  |
| --- | --- | --- | --- |
| Surfaces in isolation rooms with at least one all-day positive patient | 49 | 7 (14.3) | 0.062 |
| Surfaces in isolation rooms without any all-day positive patients | 22 | 0 (0%) |  |

Our detection of positive surface samples on one toilet ceiling-exhaust grille in a bathroom suggests that fine virus particles existed in that bathroom. However, little is known about the aerosol concentration in the bathrooms in this hospital. Deposition on exhaust grille surfaces can be a result of either long-term deposition of low-concentration particles in the air or fast deposition of high-concentration particles. It has also be confirmed that aerosols may be generated during toilet flushing.^10^ A patient’s hands can also be contaminated during his/her toilet usage. Toilet bowl and sink surface samples also tested positive in a Singaporean hospital.^11^ In addition to toilet flushing-generated aerosols, Yu et al^12^ found that SARS-CoV bio-aerosols were generated in drainage stacks after patients flushed the toilet in the 2003 Amoy Garden outbreak.

It should be noted that the toilet is a small area that is commonly shared by patients in the relatively large isolation rooms. Thus, the personal spaces of these patients overlap in the toilet area, and thus are likely to generate an additive contamination effect, and may explain why the toilet surfaces were particularly likely to be positive. Nevertheless, the duration of a patient’s stay in the toilet is brief as only mild-symptomatic patients stayed on this floor.

Additionally, the surface material or finish inside the toilets may allow a better transfer during sampling by swabs. In this context, it is known that there is generally a higher surface-touch transfer rate from smooth to rough surfaces, and most toilet surfaces are made to be smooth .^13^ We failed to detect any viruses on non-toilet object surfaces in the patients’ rooms, unlike in a 2020 study by Ong et al,^11^ who detected 13/15 (87%) positive samples in one patient’s room, and 3/5 (60%) in toilet sites, although anteroom and corridor samples were negative. It is interesting that the patient in their study had no pneumonia or diarrhoea, but that his/her stool samples were positive for SARS-CoV-2.

Among the 46 air samples, only one sample (from a corridor) was weakly positive, while all others were negative. We characterised the airflow pattern in the hospital zone using incense smoke, which revealed that there was a weak flow from all isolation rooms to their shared corridor, as shown in Figure S1. This showed that despite the continuous operation of a relatively strong exhaust fan in all isolation rooms’ bathrooms, none of these rooms had negative pressure. Thus, it is possible that leakage of aerosols from the isolation rooms to the corridor occurred. Airborne aerosols may have also been released from the protective clothing worn by healthcare workers into the environment in the multi-function room. We were not able to quantitatively measure the airflow in that zone.

Our detection of the virus in the corridor air (only weakly positive, and lower than the limit of quantification), and also on the surface of the exhaust grilles in the bathroom, suggests the possible existence of airborne viruses. There is currently no evidence for the airborne transmission of SARS-CoV-2, in contrast to the evidence for the airborne transmission of SARS-CoV in hospital wards that was presented by Li et al^14^ in 2005. Specifically, both Ong et al^11^ and Cheng et al^15^ in 2020 failed to detect any positive air samples of SARS-CoV-2, although this may have been due the fact that Ong et al^11^ sampled only 1·2 m^3^ or 1·5 m^3^ air, depending on the sampler used, while Cheng et al^15^ collected only 1 m^3^ of air. However, our single, weakly positive result was obtained from an air volume of 10 m^3^, using absorption solution as the collection medium, and no positive samples were detected when the air volume was less than 10 m^3^. Thus, it appears that the airborne concentration of SARS-CoV-2 was very low in our studied hospital.

The detection of deposited aerosols on toilet-exhaust louvres suggests the possible existence of fine airborne aerosols in the bathroom. Three possible sources exist, i.e., exhaled release from the patients when using the bathroom, toilet-generated aerosols from faeces and urine when the toilets are flushed, and import of airborne particles from the cubicles where the patients spend most of their time. Ong et al^11^ also detected SARS-CoV-2 in an air-outlet fan. Among surfaces in a patient ward, such as a bench, bedside rail, locker, bedside table, alcohol dispenser, and windowsill, Cheng et al^15^ found that only the windowsill sample (1/13) was positive for SARS-CoV-2, while the viral load of the patients was 3·3 × 10^6^ copies per mL in the pooled nasopharyngeal sample and throat swab and 5·9 × 10^6^ copies per mL in saliva.

Our data strongly imply that the toilets may be high-risk areas in hospitals with Covid-19 patients, and suggest the importance of hygiene in both private and public toilets. The strong need for hand and environmental hygiene as an intervention for Covid-19 transmission is also indicated.

There are limitations in our study. Due to the possibly strong infectivity of the new virus, there was a need for quarantine and the wearing of inconvenient full PPE by our sampling operative, which might have affected his sampling operation. Moreover, we were only permitted to access the rooms of patients with mild symptoms. The number of collected air samples was also small.

## Data Availability

Not available except those published

## Acknowledgements

The authors are grateful to many field workers and hospital staff who helped in collecting data and in providing support for our field measurements in the hospital during the most challenging period of their work.

## Authors’ contribution

The authors declare no conflict of interest.

Z. Ding, H. Qian, and Y. Li contributed equally. Y. Li, Z. Ding, H. Qian, and B. Zhu contributed to study design, hypothesis formulation, and coordination. Z. Ding and H. Qian contributed to field investigation, data analyses, and reporting. B. Xu contributed to sampling. Y. Huang contributed to patient data collection. S. Xiao to statistical analyses. X. Wu, L. Zhou and Yan Xu contributed to sampling design. W. Shao, Y. Song, S. Li contributed to sampling and patient surveys in hospital. Y. Li and Z. Ding wrote the first draft of the paper, and H. Qian, H. Yen, and T. Miao contributed to major revision. All of the other authors contributed to revision.

All of the authors approved the submitted version and have agreed to be personally accountable for their own contributions.

## Supplementary appendix

**Table S1.** Details of the patients in the wards where the sampling was conducted. The rooms containing a patient whose throat-swabs were all positive on a given day (figure below) are shown in red.

| Sampli<br>ng date | Patien<br>t no. | Bed<br>no. | Age<br>(y) | Symptom<br>onset date | No. of<br>days<br>from<br>onset to<br>samplin<br>g | Hospitalisa<br>tion date | Symptoms | CT |
| --- | --- | --- | --- | --- | --- | --- | --- | --- |
| 8 Feb | 1 | 2 | 27 | 21 Jan | 18 | 27 Jan | Fever, mild cough |  |
|  | 2 | 3 | 25 | 26 Jan | 13 | 27 Jan | Fever, mild cough |  |
|  | 3 | 16 | 65 | 4 Feb | 4 | 4 Feb | No fever | Slight patch on right lower lung |
|  | 4 | 17 | 33 | 4 Feb | 4 | 4 Feb | Fever, cough, shortness of breath |  |
|  | 5 | 18 | 38 | 25 Jan | 14 | 3 Feb | No fever, agitated, poor sleep | Multiple patchy shadows in both lungs |
|  | 6 | 31 | 52 | 25 Jan | 14 | 3 Feb | Fever | Multiple patchy shadows in both lungs |
|  | 7 | 32 | 63 | 4 Feb | 4 | 4 Feb | Fever, cough | Multiple ground- glass opacity in both lungs |
| 20 Feb<br>22 Feb | 8 | 2 | 41 | 7 Feb | 13/N.A. | 10 Feb | Fever, cough with some sputum production, chest distress, headache | Upper right lung and two lower lung lesions |
|  | 9 | 3 | 38 | 10 Feb | 10/N.A. | 10 Feb | Some diarrhoea | Two lung inflammations |
|  | 3 | 16 | 65 | 4 Feb | 16/N.A. | 4 Feb | No fever | Slight patch on right lower lung |
|  | 5 | 18 | 38 | 25 Jan | 16/N.A. | 3 Feb | No fever, agitated, poor sleep | Multiple patchy shadows in both lungs |
|  | 10 | 55 | 32 | 14 Feb | 6/8 | 16 Feb | Mild cough | Two lung inflammations |

| Sampling date | Patient no. | Bed no. | Wearing mask at sampling | Showing symptoms at sampling | Patient's nasal swab samples (+ positive; - negative) |  |  |  |  |  |
| --- | --- | --- | --- | --- | --- | --- | --- | --- | --- | --- |
| 8 Feb | 1 | 2 | Yes | No | 30 Jan (+) | 1 Feb (+) | 2 Feb (+) | 4 Feb (-) | 5 Feb (-) |  |
|  | 2 | 3 | Yes | No | 31 Jan (+) | 3 Feb (+) | 5 Feb (+) | 7 Feb (-) | 8 Feb (-) | 9 Feb (-) |
|  | 3 | 16 | Yes | No | 2 Feb (+) | 6 Feb (+) | 9 Feb (+) | 10 Feb (+) | 12 Feb (+) | 16 Feb (+) |
|  | 4 | 17 | Yes | No | 4 Feb (+) | 6 Feb (+) | 8 Feb (-) | 12 Feb (-) | 13 Feb (-) | 15 Feb (-) |
|  | 5 | 18 | Yes | No | 2 Feb (+) | 6 Feb (-) | 7 Feb (-) | 10 Feb (+) | 12 Feb (-) | 13 Feb (+) |
|  | 6 | 31 | Yes | No | 2 Feb (+) | 6 Feb (+) | 8 Feb (+) | 9 Feb (+) | 12 Feb (+) | 16 Feb (+) |
|  | 7 | 32 | Yes | No | 5 Feb (-) | 7 Feb (+) | 9 Feb (-) | 10 Feb (+) | 12 Feb (-) | 14 Feb (-) |
| 20 Feb<br>22 Feb | 8 | 2 | Yes | No | 11 Feb (+) | 13 Feb (+) | 15 Feb (-) | 17 Feb (-) | 19 Feb (-) | 21 Feb (-) |
|  | 9 | 3 | No, making phone call | No | 11 Feb (+) | 13 Feb (+) | 15 Feb (+) | 17 Feb (+) | 19 Feb (+) |  |
|  | 3 | 16 | Yes | No | 2 Feb (+) | 6 Feb (+) | 9 Feb (+) | 10 Feb (+) | 12 Feb (+) | 16 Feb (+) |
|  | 5 | 18 | Yes | No | 2 Feb (+) | 6 Feb (-) | 7 Feb (-) | 10 Feb (+) | 12 Feb (-) | 13 Feb (+) |
|  | 10 | 55 | Yes | Some coughing | 17 Feb (+) | 19 Feb (+) | 21 Feb (+) | 23 Feb (+) | 26 Feb (+) | 28 Feb (+) |

**Table S2.**
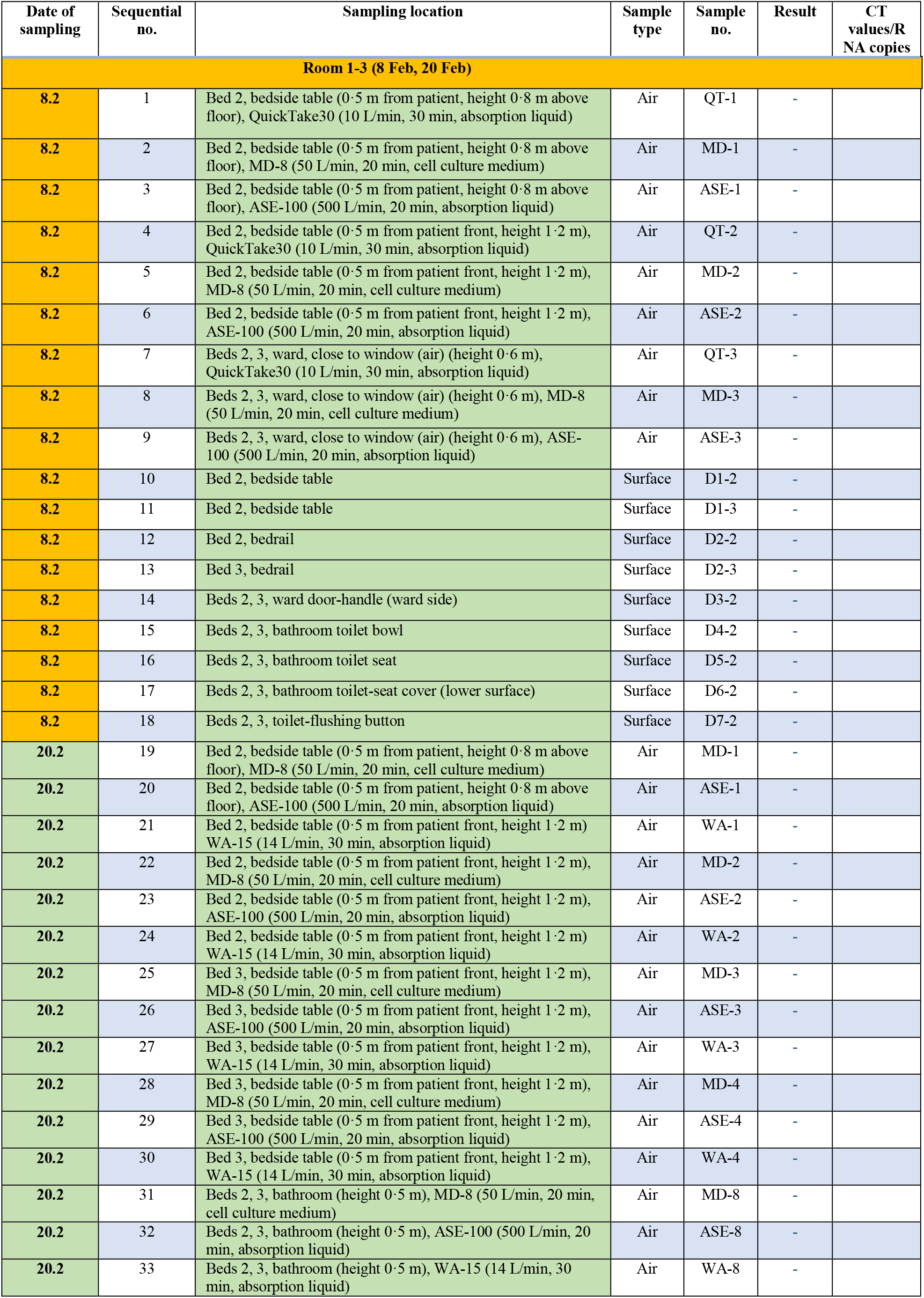
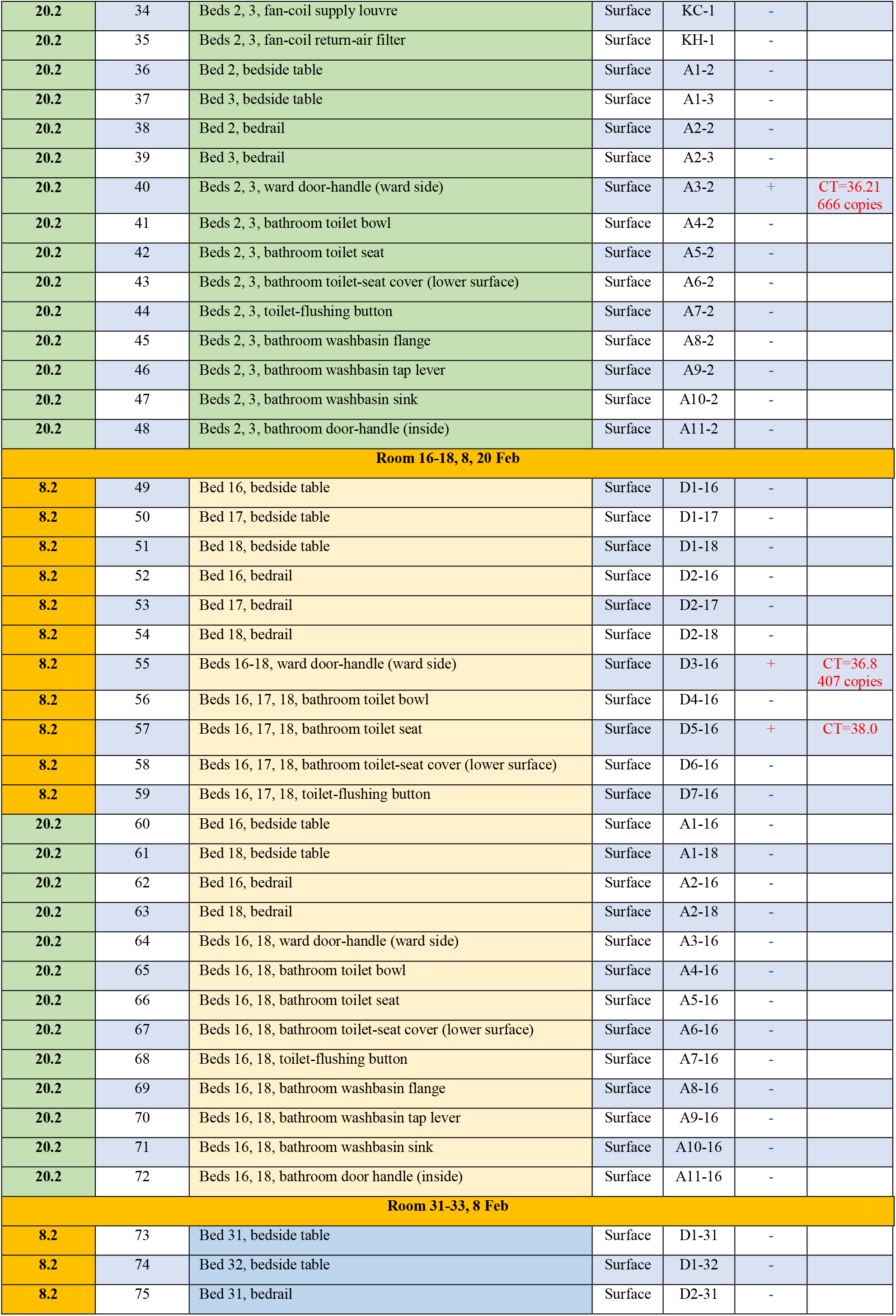
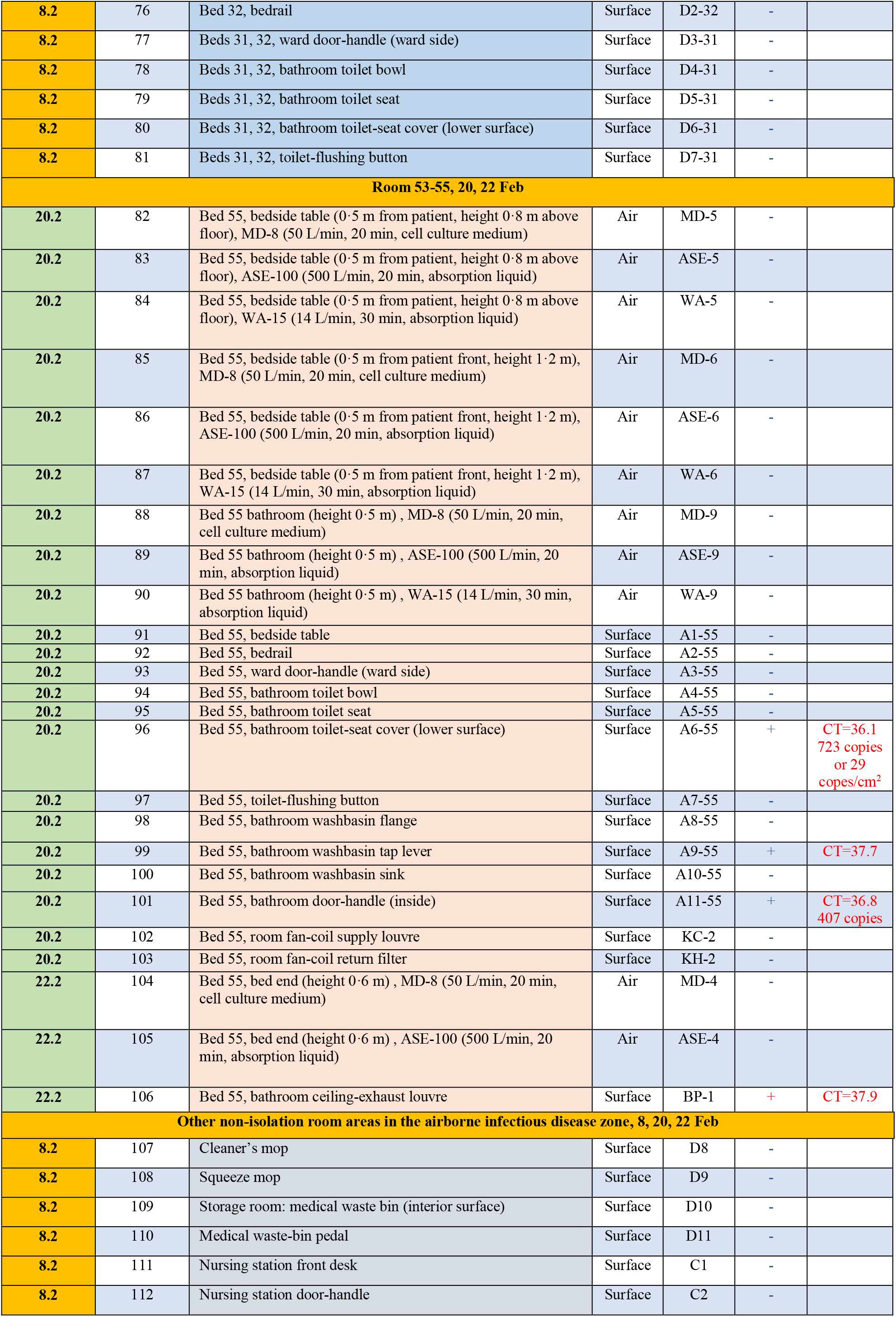
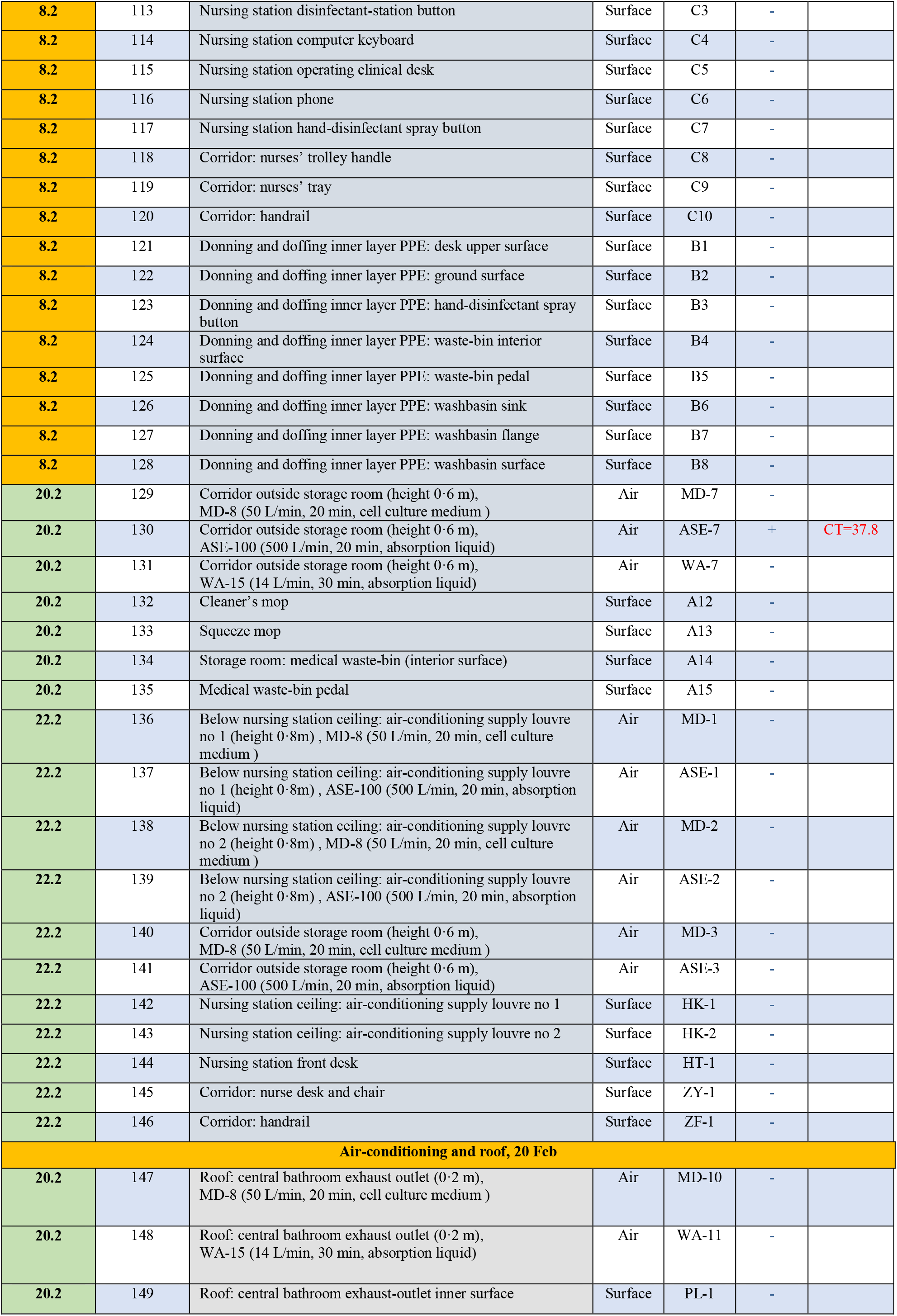
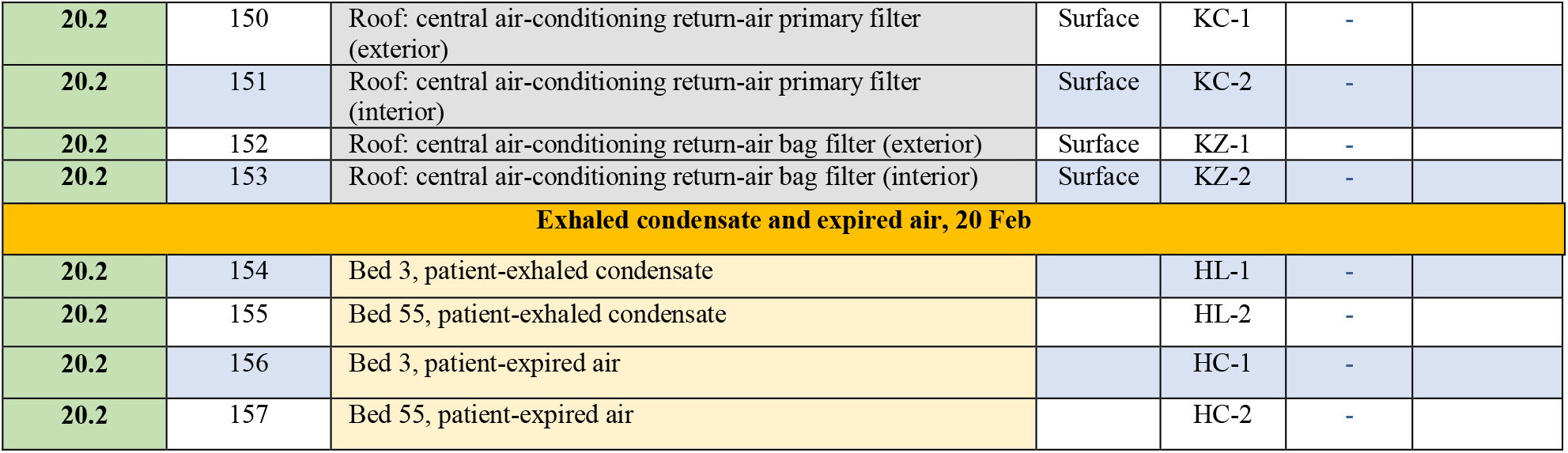
List of environmental samples Note: The indoor air-temperature on 8 Feb was 19-20°C and the relative humidity was 28-29%. The indoor air-temperature on 20 Feb was 21-23°C and the relative humidity was 27-29%. The indoor air-temperature on 22 Feb was 22-23°C and the relative humidity was 33-34%.

**Figure S1.**
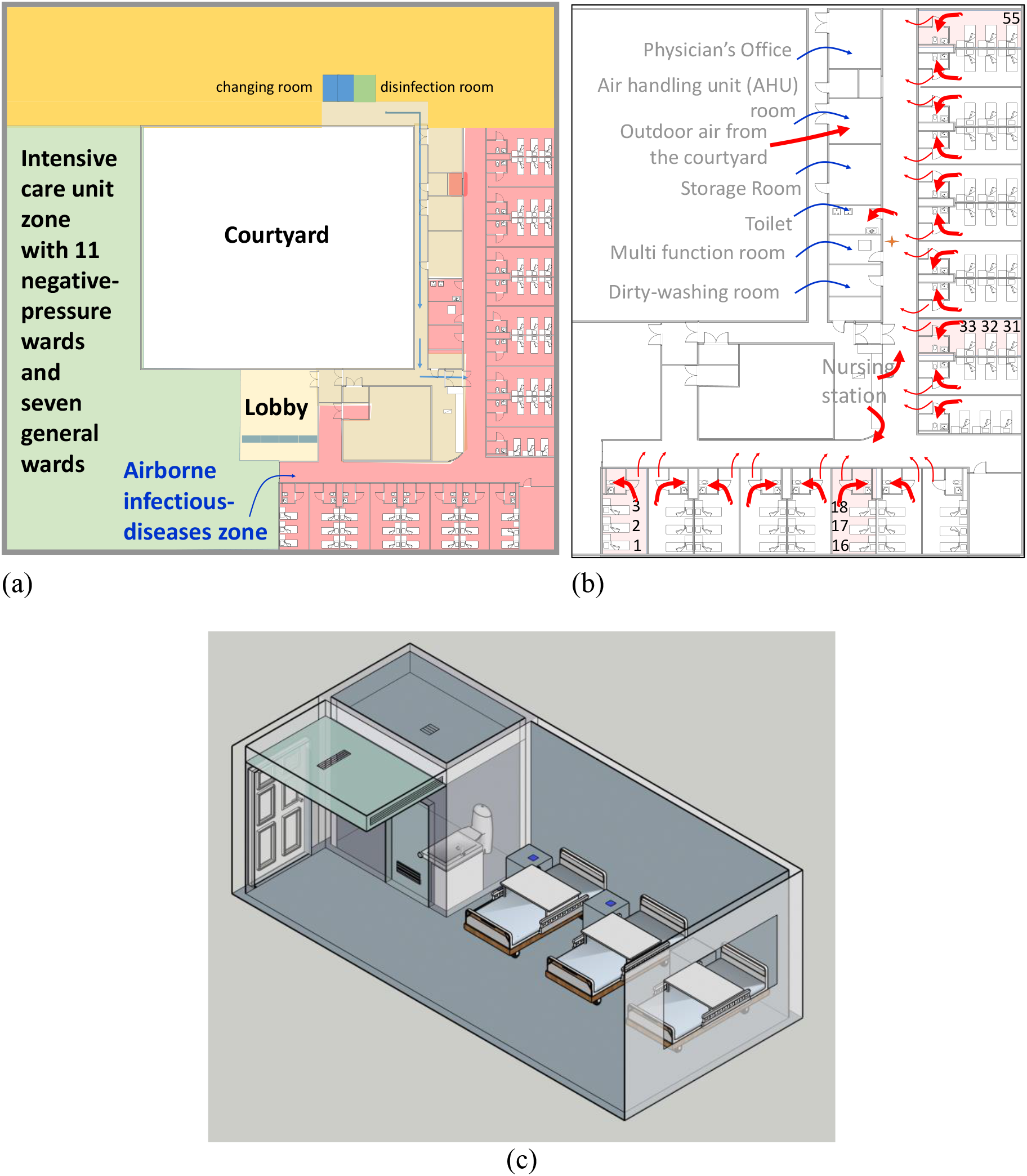
(a) Floor plan of the fifth floor of the hospital, with the airborne infectious-disease zone shown in transparent pink. (b) Floor plan of the airborne infectious-disease zone showing the location of the sampled isolation rooms (with beds 1-3; beds 16-18; beds 31-33; bed 55 shown), nursing station, and other rooms. The general airflow pattern is shown by red arrows. (c) Three-dimensional perspective view of the isolation room, showing the location of various objects.

## Notes

### Competing Interest Statement

The authors have declared no competing interest.

## References

1. WHO. 2020. Coronavirus disease 2019 (COVID-19) Situation Report – 71. https://www.who.int/emergencies/diseases/novel-coronavirus-2019/situation-reports/. Accessed on 1 April 2020.

2. WHO-China. 2020. Report of the WHO-China joint mission on coronavirus disease 2019 (Covid-19), 16–24 February 2020. https://www.who.int/docs/default-source/coronaviruse/who-china-joint-mission-on-covid-19-final-report.pdf. Accessed on 8 March 2020.

3. Zhang J, Wang S, Xue Y. Fecal specimen diagnosis 2019 Novel Coronavirus–Infected Pneumonia. Journal of medical virology. Published on 3 March 2020. https://doi.org/10.1002/jmv.25742

4. Zhang Y, Chen C, Zhu S, Shu C, Wang D, Song J, et al. Isolation of 2019-nCoV from a stool specimen of a laboratory-confirmed case of the coronavirus disease 2019 (Covid-19). China CDC Weekly 2020; 2(8): 123–4.

5. Zhang W, Du RH, Li B, Zheng XS, Yang XL, Hu B, et al. Molecular and serological investigation of 2019-nCoV infected patients: implication of multiple shedding routes. Emerging Microbes & Infections 2020; 9(1): 386–9.

6. Novel Coronavirus Pneumonia Emergency Response Epidemiology Team. Vital surveillances: the epidemiological characteristics of an outbreak of 2019 novel coronavirus diseases (COVID-19)—China, 2020. China CDC Weekly. http://weekly.chinacdc.cn/en/article/id/e53946e2-c6c4-41e9-9a9b-fea8db1a8f51. Accessed on 20 February 2020.

7. Holshue ML, DeBolt C, Lindquist S, Lofy KH, Wiesman J, Bruce H, et al. First case of 2019 novel coronavirus in the United States. New England Journal of Medicine 2020; 382:929–936.

8. Wang D, Hu B, Hu C, Zhu F, Liu X, Zhang J, et al. Clinical characteristics of 138 hospitalized patients with 2019 novel coronavirus–infected pneumonia in Wuhan, China. JAMA, 2020; 323 (11): 1061–1069. https://jamanetwork.com/journals/jama/fullarticle/2761044. 7 February 2020.

9. Peiris JSM, Chu CM, Cheng VCC, Chan KS, Hung IFN, Poon LL, et al. Clinical progression and viral load in a community outbreak of coronavirus-associated SARS pneumonia: a prospective study. Lancet 2003; 361(9371): 1767–72.

10. Gerba CP, Wallis C, Melnick JL. Microbiological hazards of household toilets: droplet production and the fate of residual organisms. Applied and Environmental Microbiology 1975; 30(2): 229–37.

11. Ong SWX, Tan YK, Chia PY, Lee TH, Ng OT, Wong MSY, Marimuthu K. Air, surface environmental, and personal protective equipment contamination by severe acute respiratory syndrome coronavirus 2 (SARS-CoV-2) from a symptomatic patient. Journal of the American Medical Association 2020; DOI: 10.1001/jama.2020.3227. Published online 4 March 2020.

12. Yu ITS, Qiu H, Tse LA, & Wong TW. Severe acute respiratory syndrome beyond Amoy Gardens: completing the incomplete legacy. Clinical Infectious Diseases 2014; 58(5): 683–6.

13. Zhao P, Chan PT, Gao Y, Lai HW, Zhang T, Li Y. Physical factors that affect microbial transfer during surface touch. Building and Environment 2019; 158: 28–38.

14. Li Y, Huang X, Yu ITS, Wong TW and Qian H. Role of air distribution in SARS transmission during the largest nosocomial outbreak in Hong Kong. Indoor Air 2005; 15:83–95.

15. Cheng VC, Wong SC, Chen JH, Yip CC, Chuang VW, Tsang OT,, et al. Escalating infection control response to the rapidly evolving epidemiology of the Coronavirus disease 2019 (COVID-19) due to SARS-CoV-2 in Hong Kong. Accepted, Infection Control & Hospital Epidemiology 2020. DOI: 10.1017/ice.2020.58.

